# Antenatal corticosteroids for pregnant women at risk of preterm labor in low- and middle-income countries: utilization and facility readiness

**DOI:** 10.1101/2024.07.31.24310863

**Authors:** Wen-Chien Yang, Catherine Arsenault, Victoria Y. Fan, Hannah H. Leslie, Fouzia Farooq, Andrea B. Pembe, Theodros Getachew, Emily R. Smith

## Abstract

**Background:** Antenatal corticosteroids (ACS) use among pregnant women with a high likelihood of preterm labor improves newborn survival. ACS adoption in low– and middle-income countries (LMICs) remains limited. Giving ACS in inadequately equipped settings could be harmful to mothers and newborns. Thus, health facilities have to demontrate readiness to administer ACS. However, the degree to which health systems are ready is unknown.

**Objective:** We assessed facility readiness to administer ACS based on the 2022 WHO recommendations on ACS use and ACS utilization.

**Methods:** The study used Service Provision Assessment surveys administered between 2013 and 2022 in nine LMICs. The primary outcome was whether facilities had ever provided ACS. We also assessed injectable corticosteroid (dexamethasone or betamethasone) availability and facility readiness to administer ACS. We used a total of 35 indicators, grouped into four readiness categories based on the WHO recommendations, to measure facility readiness.

**Findings:** Across eight countries with comparable sampling strategies, only 10.7% (median, range 6.7% – 35.2%) of facilities had ever provided ACS; one-fourth (median 25.3%, range 4.6% – 61.5%) of facilities had injectable corticosteroids available at the time of the survey; overall readiness indices were low ranging from 8.1% for Bangladesh to 32.9% for Senegal. Across four readiness categories, the readiness index was the lowest for criterion 1 (ability to assess gestational age accurately and identify a high likelihood of preterm birth) (7.3%), followed by criterion 2 (ability to identify maternal infections) (24.8%), criterion 4 (ability to provide adequate preterm care) (31.3%), and criterion 3 (ability to provide adequate childbirth care) (32.9%).

**Conclusion:** We proposed a strategy for measuring facility readiness to implement one of the most effective interventions to improve neonatal survival. Countries should operationalize readiness measurement, improve facilities readiness to deliver this life-saving intervention, and encourage ACS uptake by targeting facilities that are well-equipped.

## INTRODUCTION

Antenatal corticosteroids (ACS) use among pregnant women at risk of preterm labor is one of the most effective interventions to improve neonatal outcomes. By accelerating fetal lung maturity, ACS can reduce the risk of respiratory distress syndrome by 30% and neonatal death by 20%.(1) The World Health Organization (WHO) and other professional medical organizations recommend giving ACS to pregnant women at risk of imminent preterm labor from gestational age (GA) of 24 to 34 weeks.(2–5)

While ACS is commonly used in high-income countries, its adoption remains extremely limited in low– and middle-income countries (LMICs) despite LMICs contributing to 80% of preterm births worldwide.(6) An estimated 13.4 million newborns were born prematurly in 2020.(7) Globally, preterm births are the leading cause of neonatal death, accounting for nearly half (46%) of under-five moratlity (U5M).(8) Additionally, 75% of all neonatal deaths occur in the first week of life, with 1 million neonatal deaths happening within the first 24 hours after birth (9, 10), which motivates the interventions targteing this critical period.(11, 12) ACS has been considered “*the lowest-hanging and sweetest fruit”* as an intervention to improve preterm outcomes in LMICs.(13) However, ACS uptake in LMICs has been a subject of widespread debate.(13–18)

Two major evidence gaps remain. First, the current status of ACS use in LMICs is unknown despite some outdated coverage data. Second, the structural readiness of health facilities in LMICs to provide ACS based on international guidelines is unclear. In 2022, the WHO released its recommendations emphasizing five conditions for safe and effective administration of ACS: 1) GA can be accurately assessed, 2) there is a high likelihood of preterm birth within 7 days of starting ACS therapy, 3) there is no evidence of maternal infections, 4) adequate childbirth care is available, and 5) the preterm newborn can receive adequate care.(2)

The WHO recommendations caution that these five conditions might not be met consistently across settings because of varied capabilities, highlighting the potential harms of ACS in places lacking the capacity to meet the criteria.(2). As a potent anti-inflammatory drug, ACS suppresses immune functions. Maternal infections remain a major concern if ACS is given to vulnerable pregnant women who are ineligible for this intervention. Also, observational studies found increased neurocognitive disorders among late preterm infants (born at GA 34 to 36 weeks) who were exposed to ACS.(19–21) The balance between the benefits and risks emphasized the importance of locations in which ACS should be given. However, knowledge about facility readiness in resource-constraint settings remains scarce. In 2021, Kankaria et al. found that primary and secondary facilities in northern India were not ready to administer ACS safely, which is, to date and to our knowledge, the only study assessing facility readiness to give ACS.(22) As facility readiness is crucial in providing healthcare of good quality (23), there is an urgent need to expand evidence on ACS use in low resource countries.

This study aimed to address these knowledge gaps. The objectives were to assess ACS use, corticosteroid availability, and facility readiness to administer ACS according to the WHO criteria.

## METHODS

### Study sample

This study used data from the Service Provision Assessment (SPA), a health facility survey on service availability and quality of care.(24) We restricted our analysis to SPAs done in the past 10 years and used the latest survey available for countries with SPA data. This study included 10 surveys from 9 countries: Afghanistan 2018-2019, Bangladesh 2017-2018, Nepal 2021, Haiti 2017-2018, the Democratic Republic of Congo (DRC) 2017-2018, Ethiopia 2021-2022, Malawi 2013-2014, Senegal 2018 and 2019, and Tanzania 2014-2015. All surveys were completed before the release of the 2022 WHO recommendations but most, except Malawi 2013-2014 and Tanzania 2014-2015 surveys, were conducted after the WHO recommendations on interventions to improve preterm birth outcomes in 2015, which contained guidelines on ACS use.(25) The sampling strategies varied based on country needs.(26–35) The majority of surveys adopted stratified random sampling strategies to obtain a national representative sample in the country.(26–28, 31, 32) Malawi 2013-2014 and Haiti 2017-2018 SPAs were national census.(33, 35) Unlike other surveys, Afghanistan 2018-2019 mainly sampled urban hospitals.(34) Senegal implemented continuous SPA over a consecutive 5 years to survey all health facilities.(29, 30, 36) In order to have similar sample sizes across countries, we merged two years (2018 and 2019) of data from Senegal **(Supplemental Table 1)**. Our study used data from two core instruments (out of five) of the SPA survey questionnaire: facility inventory and health worker interviews. In each country, only facilities that provided antenatal care (ANC), performed normal deliveries, and/or performed Cesarean deliveries were included.

### Measures

Our primary outcome was ACS utilization, defined as facilities that had ever provided ACS to pregnant women. We focused on two secondary outcomes: corticosteroid availability (injectable dexamethasone or betamethasone) and facility structural readiness. Most countries did not survey corticosteroid availability (except for Afghanistan 2018-2019) within the maternal and child health care sections in SPA surveys. We reported the availability of injectable corticosteroids that were surveyed in the section on medicines for non-communicable diseases.

We assessed facility readiness to provide ACS in accordance with the 2022 WHO recommendations on ACS use. We identified 35 indicators from the SPA questionnaire, which were grouped into four readiness categories based on the five WHO criteria (collapsing criterion 1 and criterion 2) as outlined in **Supplemental Table 2**. The first category focused on the facility’s ability to assess GA accurately and to identify pregnant women with a high likelihood of preterm labor; we included only one indicator for this category (presence of a functional ultrasound). The second category included 4 indicators and covered the facility’s ability to identify maternal infections. The third and fourth categories included 13 and 17 indicators and assessed the facility’s readiness to provide adequate childbirth care and preterm newborn care, respectively. For each category, readiness indices were calculated by dividing the number of indicators available by the total number of indicators assessed, where higher percentages indicating higher readiness. An overall readiness index was calculated by averaging the readiness indices from four categories. This approach was chosen based on previous studies on facility readiness to implement health improvement interventions.(37, 38)

### Statistical analysis

First, we presented descriptive statistics of ACS utilization, corticosteroid availability, and facility readiness indices, taking into account survey sampling weights. Results were stratified by three facility levels. Level 1 facilities were those that only provided ANC and did not conduct deliveries; ANC was defined as the care that pregnant women receive before birth, including risk identification, prevention, and management of pregnancy related health conditions, education, and health promotion.(39) Level 2 facilities performed normal deliveries, but not Cesarean deliveries, and level 3 facilities performed Cesarean deliveries. Because Afghanistan mainly sampled hospitals, which substantially differed from other countries, it was excluded from the summary descriptive analysis of our outcomes of interest and included only in analyses by facility levels.

Second, we assessed the relationships between ACS use and, respectively, ACS availability, and overall readiness indices at sub-national levels (e.g. regions in Ethiopia, provinces in DRC). This was driven by the hypothesis that facilities without corticosteroids or with low readiness might refer patients in need of ACS to facilities in geographic proximity. We averaged each of the three measures (ACS use, corticosteroid availability, and readiness index) for all facilities in each sub-national region. Lastly, we examined the difference in the overall readiness index between facilities that had ever and never used ACS by country. All analyses were performed using R.

## RESULTS

This study included a total of 8669 facilities from ten surveys in nine countries. All surveys had a high response rate (median 94.9%) ranging from 88.8% in Afghanistan to 99.0% in Tanzania **(Supplemental Table 3)**.

The majority (88.9%, median; range 66.1% – 98.8%) of facilities provided maternal health care services and were included in our analysis **(Supplemental Table 3)**. The median sample size was 929 facilities (median; range 108 –1500) **(Table 1)**. Among eight countries (excluding Afghanistan), 22.6% (median; range 7.1% – 88.7%) of facilities were urban. The proportion of facilities of different levels varied across countries. Across eight countries, one third (median 32%) of the facilities were level 1 facilities that provided antenatal care, with Bangladesh having the highest proportion (76.2%) of level 1 facilities and the DRC having the lowest proportion (1.9%). The proportion of level 2 facilities was 59.4% (median) with Tanzania having the highest proportion (83.3%) and Bangladesh with the lowest proportion (19.5%). Only 6.2% (median) of facilities were level 3 facilities that performed Cesarean deliveries, with 26.5% of facilities in DRC and 2.7% in Ethiopia were level 3 **(Table 1)**.

**Table 1.** Characteristics of surveys and facilities included in the study.

|  |  |  |  |  |  |  |  |
| --- | --- | --- | --- | --- | --- | --- | --- |
| Country | Survey year | Number of facilities surveyed<sup>1</sup> | Number of facilities included in the analysis<sup>2</sup> | Urban location | Facility type |  |  |
|  |  |  |  |  | Level 1:<br>Facilities that provide antenatal care | Level 2:<br>Facilities that perform normal delivery | Level 3:<br>Facilities that perform Cesarean section |
|  |  | N = 9793<br>Median 1158<br>(range 142 - 1576) | N = 8669<br>Median 929<br>(range 108 - 1500) | Median 22.6%<sup>3</sup><br>(range 7.1% - 88.7%)<sup>3</sup> | Median 32.0%<sup>3</sup><br>(range 1.9% - 76.2%)<sup>3</sup> | Median 59.4%<sup>3</sup><br>(range 19.5% - 83.3%)<sup>3</sup> | Median 6.2%<sup>3</sup><br>(range 2.7% - 26.5%)<sup>3</sup> |
| South Asia |  |  |  |  |  |  |  |
| Afghanistan | 2018-2019 | 142 | 108 | 107 (99.4%) | 7 (4.1%) | 10 (10.1%) | 91 (85.9%) |
| Bangladesh | 2017-2018 | 1524 | 1498 | 379 (7.1%) | 676 (76.2%) | 551 (19.5%) | 271 (4.4%) |
| Nepal | 2021 | 1576 | 1500 | 961 (53.2%) | 693 (47.5%) | 565 (47.1%) | 242 (5.3%) |
| Caribbean |  |  |  |  |  |  |  |
| Haiti | 2017-2018 | 1007 | 929 | 340 (36.5%) | 567 (61.0%) | 255 (27.5%) | 107 (11.5%) |
| Sub-Saharan Africa |  |  |  |  |  |  |  |
| DRC | 2017-2018 | 1380 | 1364 | 297 (22.0%) | 10 (1.9%) | 511 (71.6%) | 843 (26.5%) |
| Ethiopia | 2021-2022 | 1158 | 911 | 455 (18.2%) | 261 (75.0%) | 309 (22.3%) | 341 (2.7%) |
| Malawi | 2013-2014 | 977 | 645 | 119 (18.6%) | 103 (16.5%) | 471 (72.6%) | 71 (10.9%) |
| Senegal | 2018 and 2019 | 841 | 658 | 594 (88.7%) | 65 (14.3%) | 531 (78.6%) | 62 (7.1%) |
| Tanzania | 2014-2015 | 1188 | 1056 | 325 (19.1%) | 104 (11.5%) | 681 (83.3%) | 271 (5.2%) |

<sup>1</sup> A total of 625 facilities that were sampled had a sampling weight of zero, indicating non-response, refusal, or closure. Those facilities were not surveyed and were excluded.
<sup>2</sup> Only facilities that provided either antenatal care, performed normal delivery or Cesarean section were included in the analysis.
<sup>3</sup> The median and range were obtained across eight countries excluding Afghanistan because its survey used a remarkably different sampling strategy.

### ACS utilization

ACS was underutilized across all countries. Excluding Afghanistan, median ACS use was 10.7%, ranging from 6.7% of facilities in Bangladesh to 35.2% of facilities in the DRC having ever administered ACS **(Figure 1a)**. In Afghanistan, 68.7% of facilities had administered ACS at the time of the survey. We did not find higher ACS use in surveys that were done in more recently. While none of the level 1 facilities had provided ACS, ACS use increased by facility level **(Figure 1b)**. Within facility level across 9 countries, 21.6% (median, range 5.0% – 26.9%) of level 2 facilities and 76.3% (median, range 52.7% – 90.3%) of level 3 facilities had provided ACS.

**Figure 1a.**
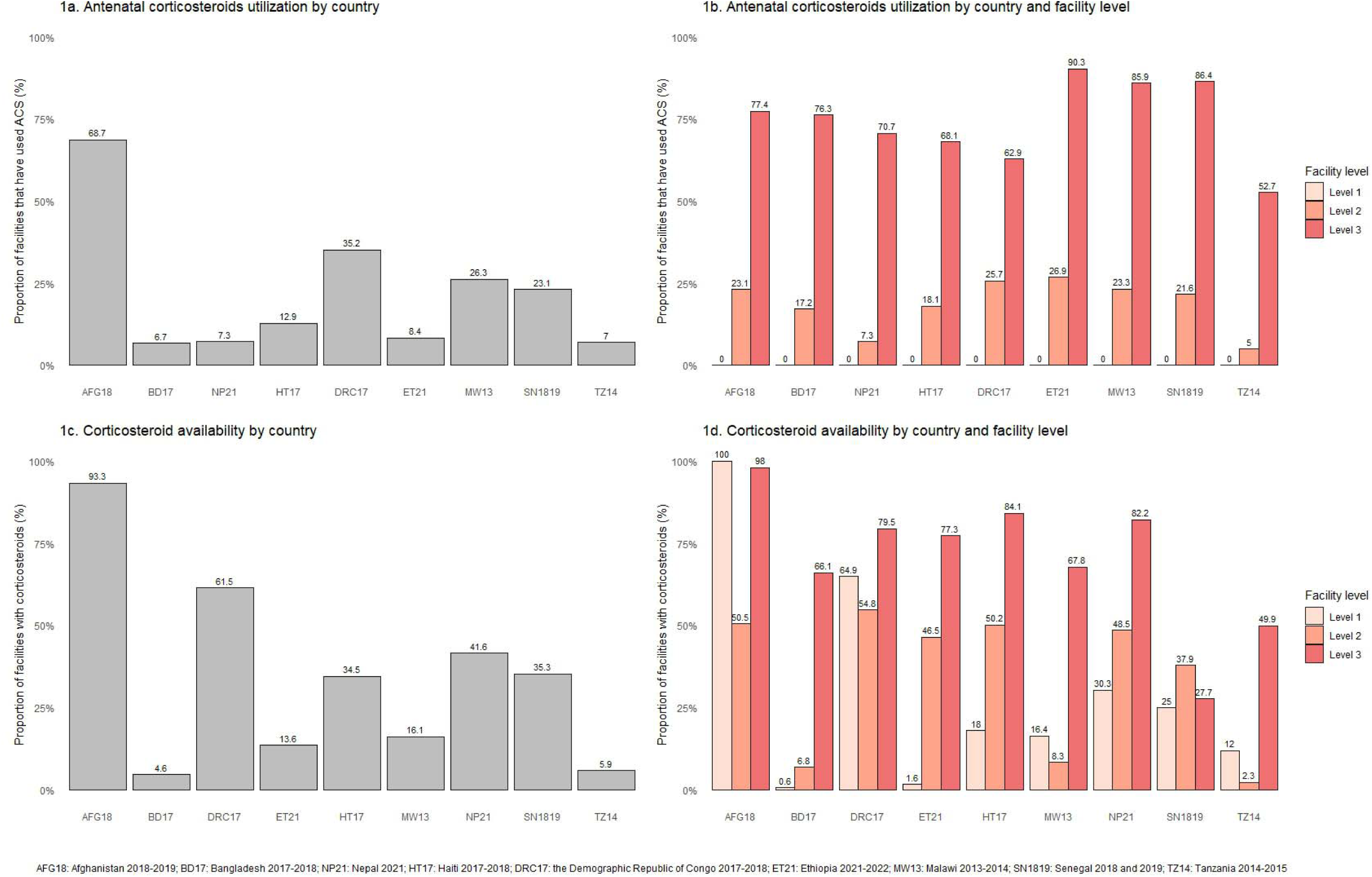
Utilization of antenatal corticosteroids by country. **Figure 1b**. Utilization of antenatal corticosteroids by country and facility level **Figure 1c**. Availability of at least one valid corticosteroid by country **Figure 1d**. Availability of at least one valid corticosteroid by country and facility level

### ACS availability

Corticosteroid availability was limited. Across eight countries, a fourth of the facilities (median 25.3%, range 4.6% – 61.5%) had at least one valid corticosteroid (injectable betamethasone or dexamethasone) available at the time of the survey, while 93.3% of facilities in Afghanistan had corticosteroids available **(Figure 1c)**. Corticosteroid availability generally increased by facility level within each country **(Figure 1d)**. Also, gaps between corticosteroid availability and ACS utilization existed. For example, 48.5% of level 2 facilities in Nepal had corticosteroid available at the time of survey but only 7.3% had ever used it **(Supplemental Figure 5)**.

### Facility readiness

Readiness indices were low **(Figure 2)**. Overall, only 22% of the facilities in the sample had an overall readiness index above 50%. Other than Afghanistan, overall readiness indices were low among the eight countries ranging from 8.1% in Bangladesh to 32.9% in Senegal. Afghanistan had an overall readiness index of 57.7%. We did not see higher overall readiness indices in surveys done in more recent years. Also, overall readiness indices increased by facility level. Across four readiness categories among 8 countries, facilities performed the worst in the ability to assess GA accurately, with a readiness index of 7.3% (median), followed by the ability to identify maternal infection (median, 24.8%), provide adequate preterm care (median 31.3%), and provide adequate childbirth care (median 32.9%). When delving into the details of specific indicators among the 35 ones used, we found very limited availability for respiratory support-related equipment across countries in the readiness category of adequate preterm newborn care **(Supplemental Table 4, Supplemental Figures 1-4)**.

**Figure 2.**
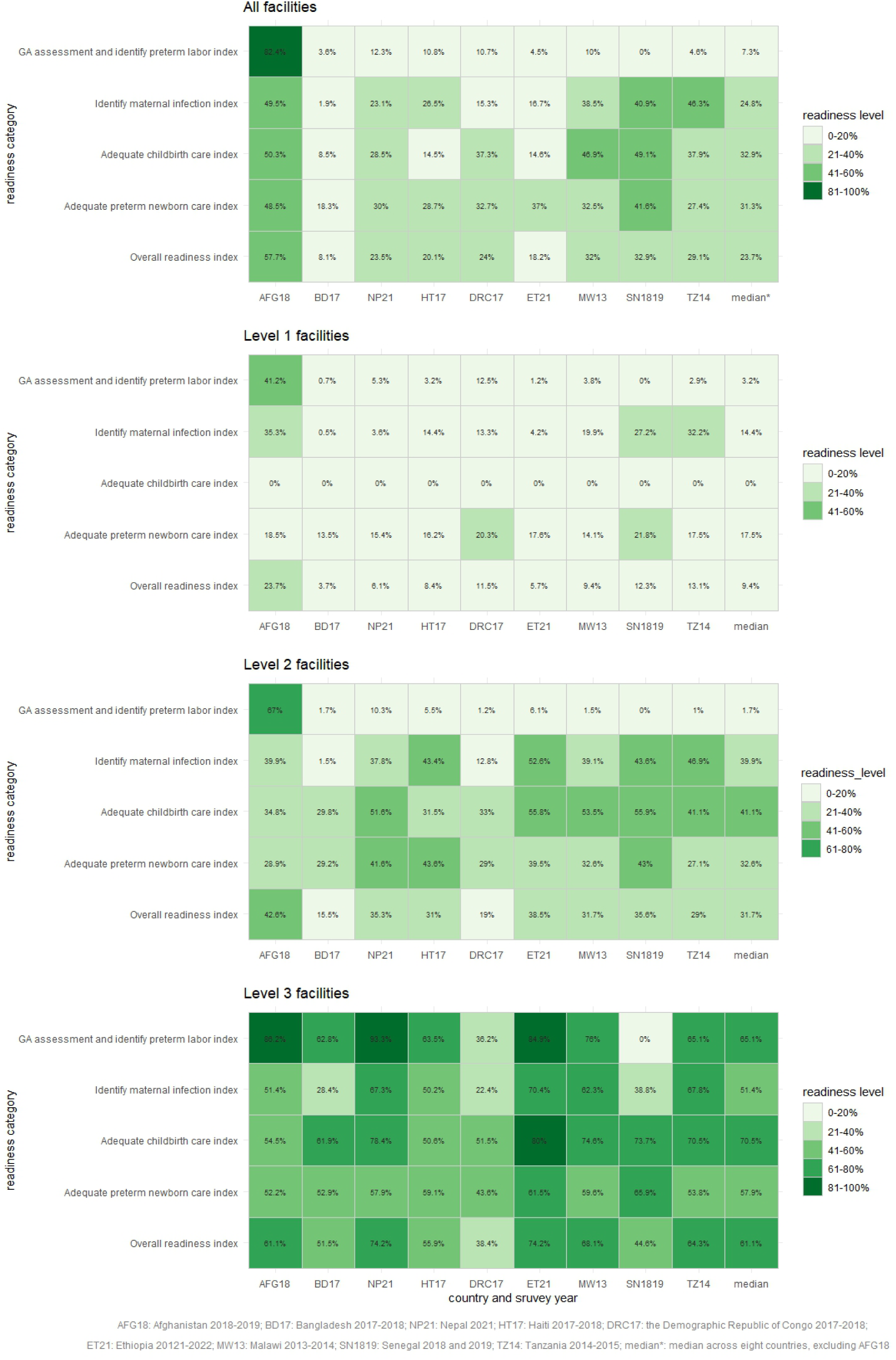
Facility readiness by country, readiness category, and facility level

### ACS use, availability, and readiness at sub-national levels

At the sub-national level, positive associations were observed between corticosteroid availability and ACS use as well as between average readiness and ACS use **(Figures 3a and 3b)**. A few Ethiopian regions (Dire Dawa and Harari) had an average overall readiness index above 50% but low ACS use (<25%). In contrast, some regions of the DRC (Kasaï Central, Kongo Central, Kinshasa, and Haut-Katanga) had low overall readiness indices (<50%), but ACS use greater than 50% **(Figure 3b)**.

**Figure 3a.**
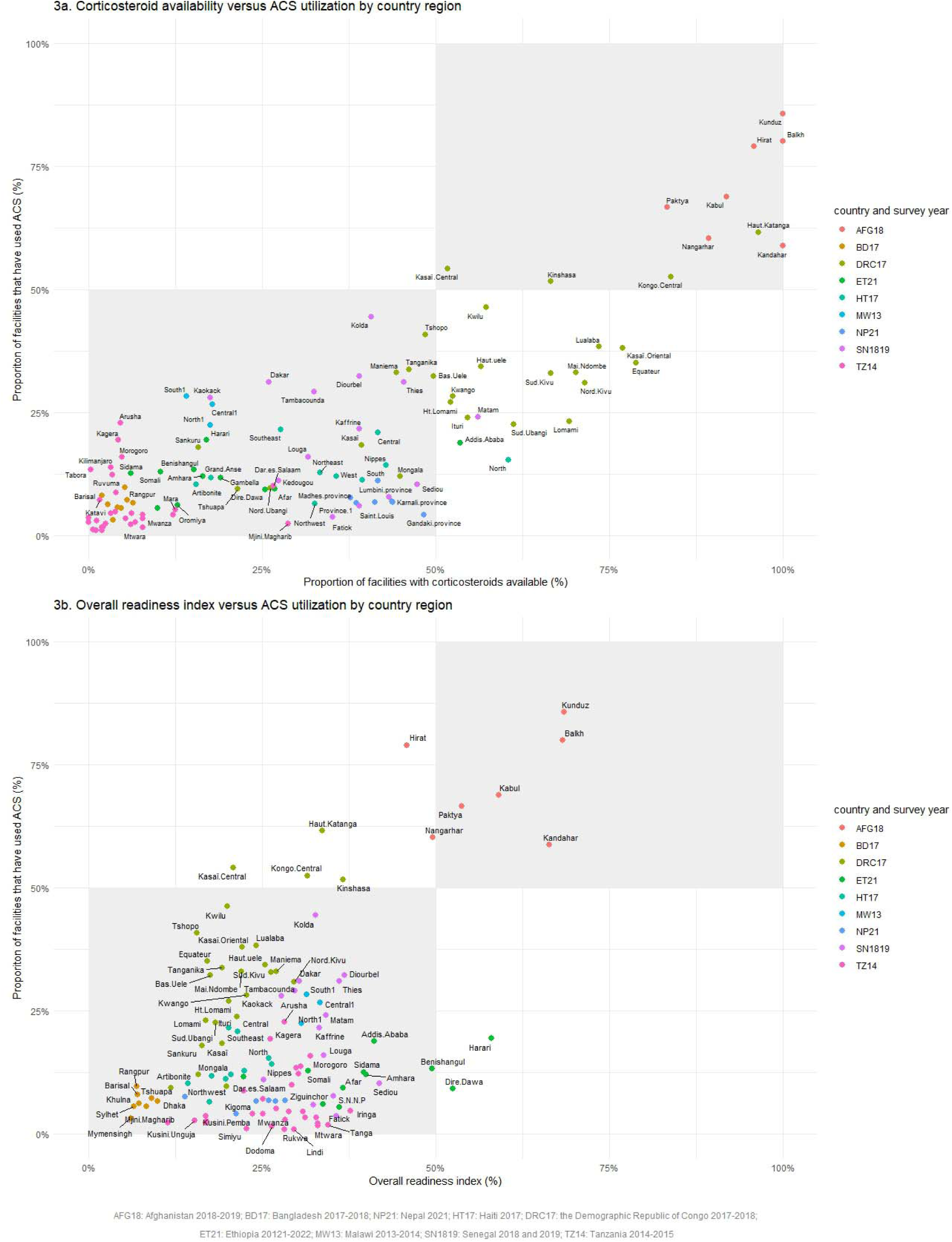
Corticosteroid availability versus ACS utilization by region. **Figure 3b**. Overall readiness indices versus ACS utilization by region

At the facility level, overall readiness indices differed between facilities that had ever and never provided ACS within each country **(Supplemental Figures 6-8)**. In Afghanistan, Bangladesh, DRC, Senegal, and Tanzania, facilities that had utilized ACS had higher overall readiness indices compared to facilities that had never used ACS. On the contrary, the results were reversed for Nepal, Haiti, Ethiopia, and Malawi.

## DISCUSSION

This study assessed antenatal corticosteroid use, corticosteroid availability and structural readiness to administer the drug adequately based on international recommendations among 8,669 health facilities from nine resource-constrained countries. We had three major findings: 1) antenatal corticosteroids were substantially underused, 2) corticosteroid availability was limited, and 3) facilities in these countries had low levels of readiness.

We found limited ACS use with only one out of ten health facilities having ever administered ACS. However, it is challenging to compare our results with existing literature as previous work mostly measured ACS coverage among pregnant women who delivered preterm infants.(40–45) A 2011 WHO maternal and newborn health survey for 29 countries found 54% of women who gave birth at GA 26 to 34 weeks were given ACS, with the lowest use in Afghanistan (16%), Nepal (20%), and DRC (16%).(41) Another analysis using the data from the 2015 Antenatal Corticosteroids Trial (ACT trial) showed an overall low ACS use in control clusters; in Kenya, only 3.8 % of pregnant women of infants born with birthweight less than the 5^th^ percentile, a proxy for preterm births, received ACS.(40) Our findings adds value in understanding the status of ACS use at the facility level. Also, we found that only one third of the facilities in the sample had either injectable dexamethasone or betamethasone. Dexamethasone is on the WHO List of Essential Medicines, but limited availability remains a major barrier to ACS use.(46) Strategies to improve ACS coverage need to ensure drug availability.

The location where ACS is given is crucial to ensure safe and effective use. Two landmark studies on the effects of ACS in LMICs presented conflicting results, highlighting the importance of settings in which ACS is given.(47, 48) The 2015 ACT trial, a cluster randomized trial of a multifaceted intervention to promote ACS use in six LMICs unexpectedly found increased neonatal deaths and suspected maternal infections among intervention clusters.(47) On the contrary, the WHO Antenatal Corticosteroids for Improving Outcomes in Preterm Newborns Trial (ACTION-I trial) in 2020, an RCT in five LMICs, showed ACS reduced neonatal deaths without increasing maternal infections.(48) These contradictory findings can partially be explained by the different settings for the two trials; the ACT trial was done in all levels of care including clinics and primary care centers, whereas the ACTION-I trials included secondary or tertiary hospitals.(48) The drastically different findings emphasized the importance of locations – or more precisely, the readiness of facilities – in providing ACS. Our study offered a strategy for measuring facility readiness to implement this life-saving intervention. However, recommendations on what levels of facilities should give ACS need to be made carefully.

Assuming essential equipment, medicines and trained staff are available, some level 2 and level 3 facilities with high readiness that have never provided ACS should be targeted for the expansion of ACS use. Also, our sub-national level analyses showed that some provinces in DRC with low overall readiness (<50%) frequently administered ACS. Policymakers should ensure that ACS is delivered in well-equipped settings.

Another critical issue centers around the different aspects of readiness to ensure safe and effective ACS use. Our readiness index was developed based on the WHO criteria. However, it is debatable, first, whether some criteria could be met across facility levels, and second, whether facilities need to meet *all* criteria to safely and effectively administer ACS. For example, ultrasound examination in early pregnancy is the gold standard for GA assessment. However, we found that only 7.3% (median across eight countries, range 0.2% to 12.3%; 82.4% for Afghanistan) of maternal care facilities had a functional ultrasound. Access to ultrasound for GA dating remains a major barrier to proper ACS use despite its increasing use in obstetric care in LMICs.(17, 49) In this case, GA dating should occur early in pregnancy, while ACS use occurs later in pregnancy. Thus, the decision to administer ACS should not be based on the ultrasound availability in place, but the availability of accurate GA assessment (which may come from care obtained at another facility). In addition, regarding the WHO recommendations for adequate preterm newborn care, one important element is non-invasive respiratory support, such as continuous positive airway pressure (CPAP). However, this recommendation might exclude most preterm infants who might benefit from ACS use because they usually do not have access to respiratory support in LMICs.(50) A similar concern applies to adequate childbirth care, referring to nine signal functions of Comprehensive Emergency Obstetric and Newborn Care (CEmONC), including blood transfusions and Cesarean sections. Again, it is debatable whether a facility can give ACS only when it can do blood transfusions.

Our study had several strengths. To our knowledge, this is the first study to assess ACS use, availability and structural readiness to give ACS at the facility level. Also, we included data from multiple countries to understand the landscape of ACS use and identify policy directions in areas with a high burden of preterm births. Nonetheless, our study has a few limitations. First, sampling strategies and survey years varied across countries spanning 9 years. All surveys were done prior to the release of the 2022 WHO recommendations while most of them were completed after the release of 2015 WHO recommendations on interventions to improve preterm birth outcomes that contained guidelines on ACS utilization.(2, 25) Also, countries have different national guidelines regarding ACS use. One policy analysis on ACS use in Africa found ACS could be given in lower levels of care before referral in Ethiopia and Tanzania, but DRC and Malawi only allowed ACS use in hospitals.(51) Hence, we caution careful interpretations when comparing across countries. Secondly, some readiness indicators served as proxies. For instance, SPA surveys do not assess the availability of corticosteroids and ultrasound in the maternal care section. Instead, corticosteroid availability is surveyed in the section for medicines for non-communicable diseases, and ultrasound availability is assessed within the facility in general. We also used the availability of rapid diagnostic tests for HIV and syphilis as a proxy to estimate facilities’ ability to detect maternal infections. This approach might overestimate or underestimate true availability or readiness. Thirdly, we were unable to draw correlations between ACS use or facility readiness with preterm health outcomes because SPAs do not have patient-level data for newborns.

Antenatal corticosteroid use has gained tremendous international attention as its population-level health benefits could be profound.(12, 52–55) ACS should not be used as a “just-in-case” intervention.(56) It should be given to the right people (pregnant women at risk of imminent preterm labor and without maternal infections), at the right time (the specified GA window), and in the right place (settings that are properly equipped and ready to provide quality maternal and newborn care). Future research and programs should operationalize readiness measurement and assess facility readiness for safe and effective ACS use to enhance readiness and encourage ACS uptake among well-prepared facilities.

### Disclosure of relationships and activities

All authors have completed the ICMJE uniform disclosure form at www.icmje.org/coi_disclosure.pdf and declare no support from any organization for submitted work; no financial relationships with any organization that might have an interest in the submitted work in the previous three years; no other relationships or activities that could appear to have influenced the submitted work.

## Supporting information

S1 Supplementary file

## Data Availability

All data used in the present study are available on the DHS SPA website.

https://dhsprogram.com/data/

## Supplementary file

**Supplemental Table 1.** Sampling strategies of included SPA surveys.

| **Survey** | **Sampling strategy** |
| --- | --- |
| Afghanistan 2018-2019 | The survey focused on major public and private hospitals in the main seven provinces (including Nangarhar, Paktya, Kunduz, Balkh, Kandahar, and Herat, and Kabul) in the country. Among six provinces excluding Kabul, all 12 public hospitals, 37 private hospitals, and 52 private clinics were included. In Kabul, all 26 public and 20 private hospitals were surveyed, but a randomly selected sample of 13 private clinics (out of 84) were included. |
| Bangladesh 2017-2018 | The survey adopted a stratified random sampling strategy for 1,600 facilities to obtain nationally representative data for different facility types, different facility management authorities, and each of the eight divisions (Barisal, Chittagong, Dhaka, Khulna, Mymensingh, Rajshahi, Rangpur, and Sylhet) of the country. |
| Nepal 2021 | The survey adopted a stratified random sampling strategy for 1,633 health facilities through an equal probability systematic sampling. All government hospitals, all non-government hospitals with at least one bed, and all non-government hospitals in the provinces of Karnali and Sudurpashchim were included because of their small numbers. Also, all primary health care centers (PHCCs) and stand-alone HIV testing and counseling centers (stand-alone HTC) were included. |
| Haiti 2017-2018 | The survey was a national census of all operational health facilities in the country during the survey period of 2017 to 2018. |
| Demographic Republic of Congo 2017-2018 | The survey adopted a probability sampling strategy from 12,059 health facilities in the country, excluding health posts. On average, approximately 50 facilities were selected for each of the 26 provinces, reaching a sample of 1,380 facilities. |
| Ethiopia 2021-2022 | The survey adopted a stratified random sampling strategy reaching a sample of 1,407 health facilities via an unequal probability systematic sampling. Within each region, stratification was achieved by facility type. All public and private hospitals were included because of their small numbers and crucial roles in the country's health system. Health centers were sampled but all health centers in Dire Dawa and Harari were included. Clinics were sampled with all higher clinics included and all clinics in Harari included. Health posts were sampled. |
| Malawi 2013-2014 | The survey was a national census of all formal-sector health facilities, including public, private and other managing authority types, in the country during the survey period between 2013 to 2014. |
| Senegal 2018 and 2019 | Senegal implemented a continuous SPA, which was designed to continuously collect data on health facilities over a period of 5 years from 2012 to 2017, aiming to survey all health facilities in the country. |
| Tanzania 2014-2015 | The survey was a sample of formal-sector health facilities, including public and private, during the survey period between 2014 to 2015. The sample was selected to obtain national representative data by facility type, managing authority type, and regions. |

**Supplemental Table 2.**
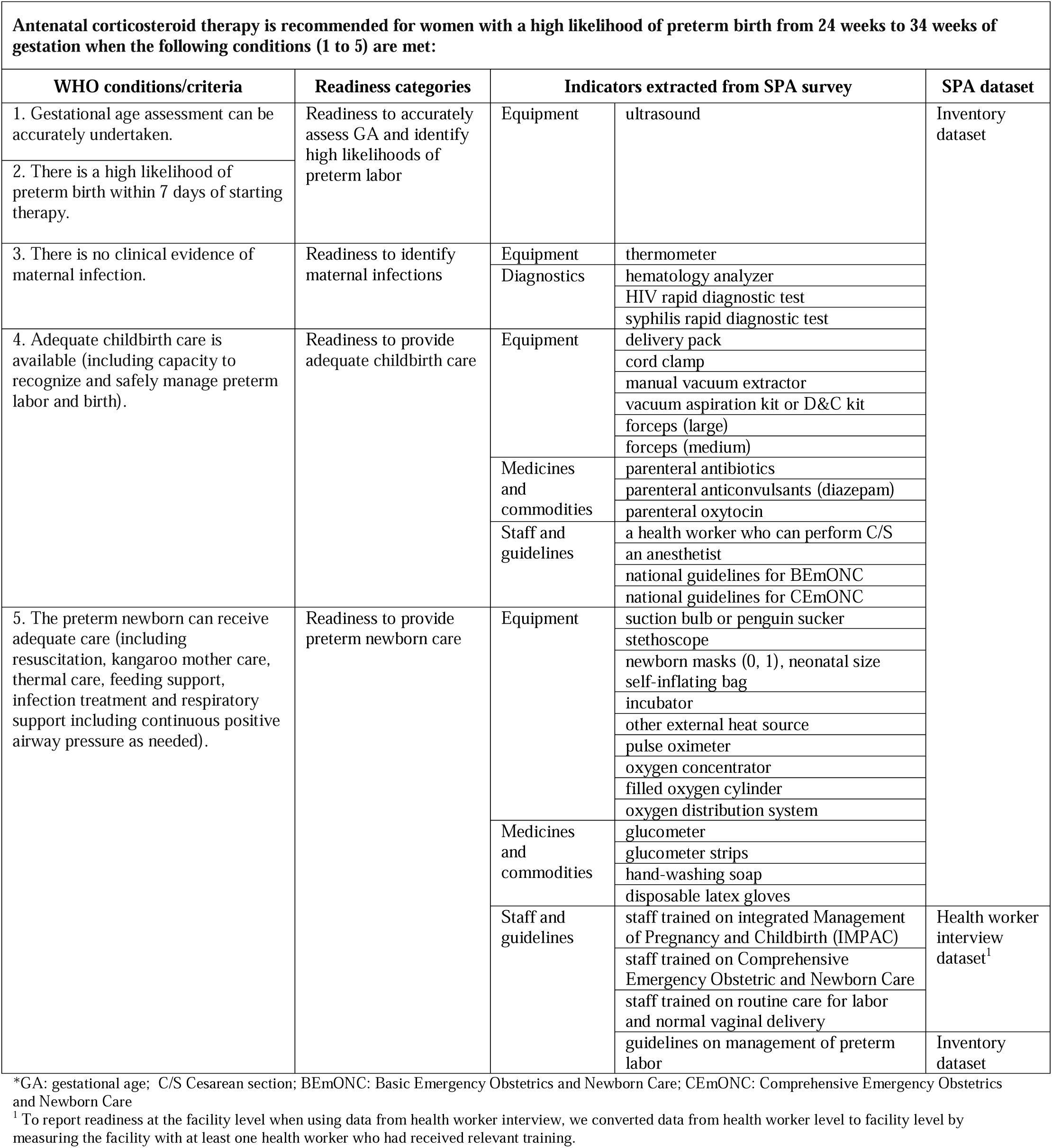
The indicators used to assess facility readiness based on the 2022 WHO commendations on ACS use.

**Supplemental Table 3.**
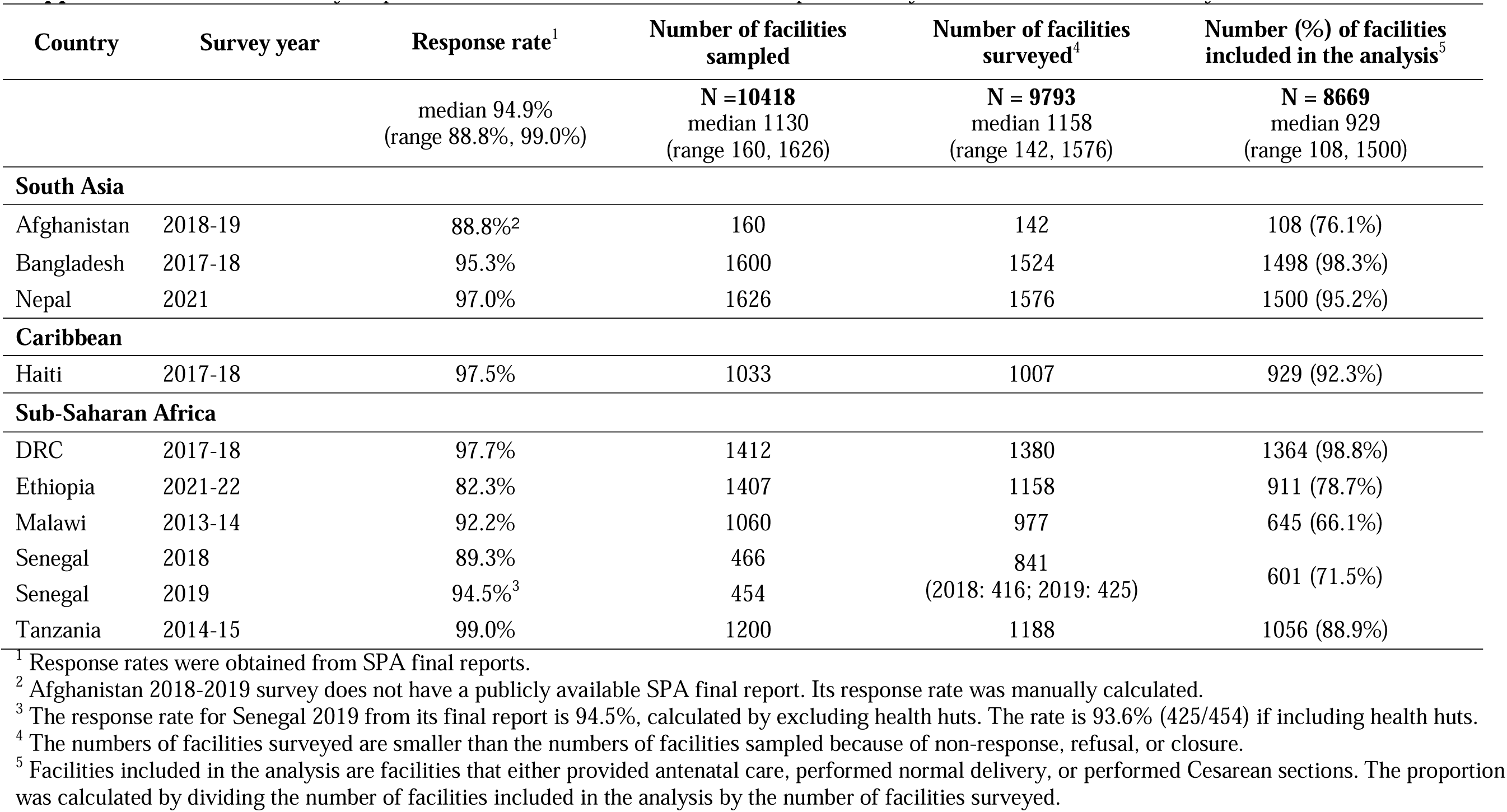
Survey response rates and number of facilities sampled, surveyed, and included in the analysis.

**Supplemental Table 4.**
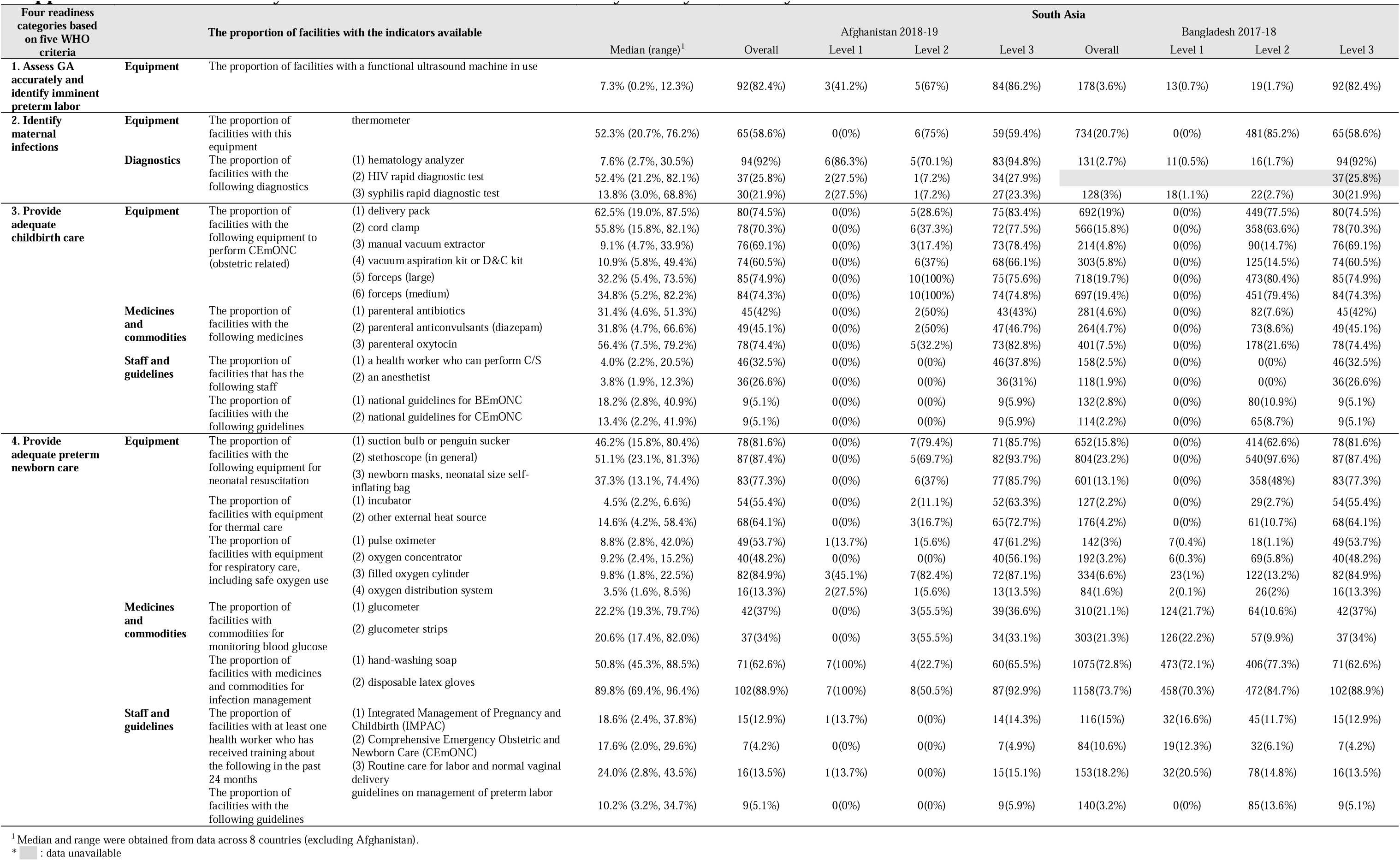
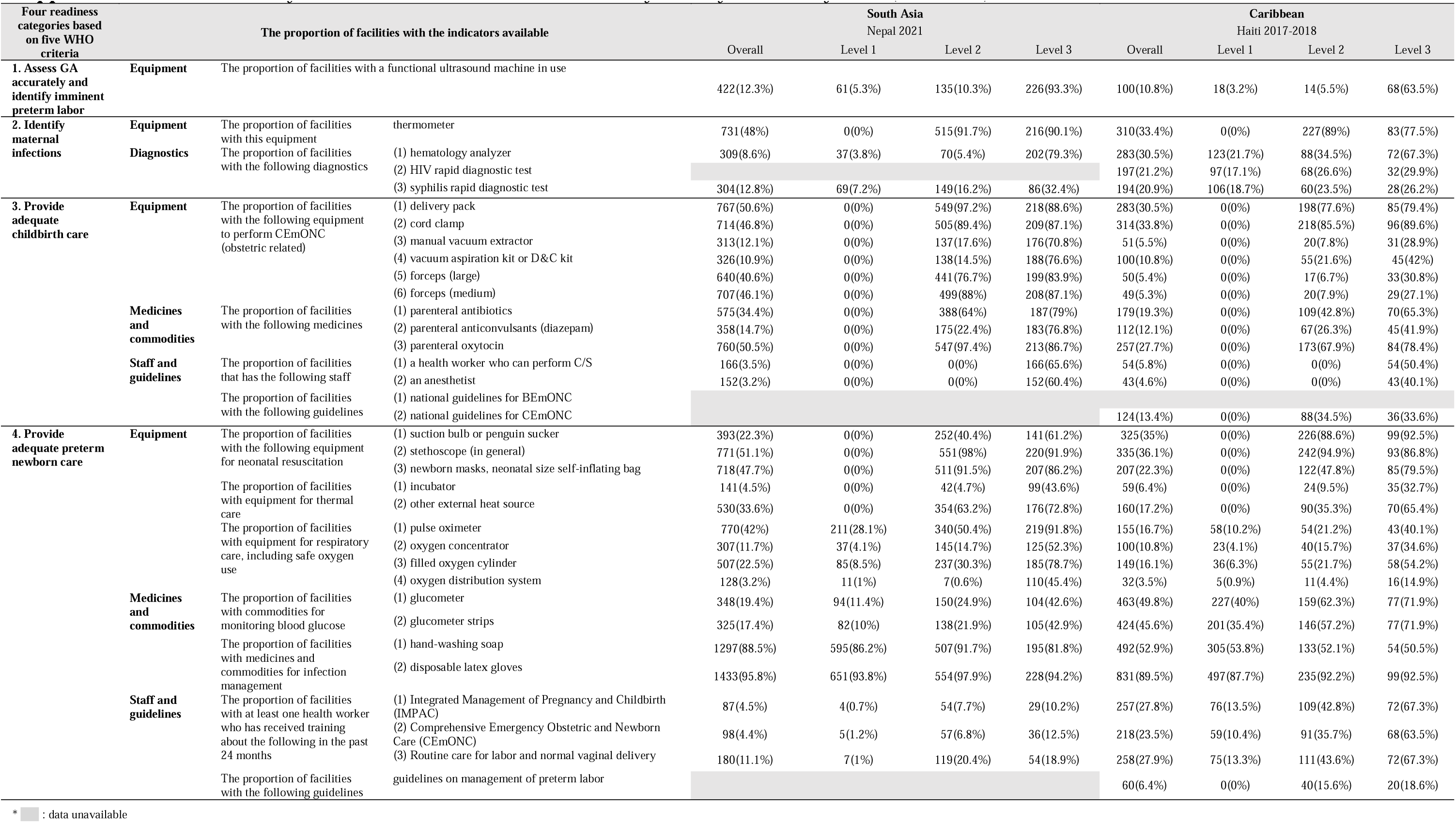
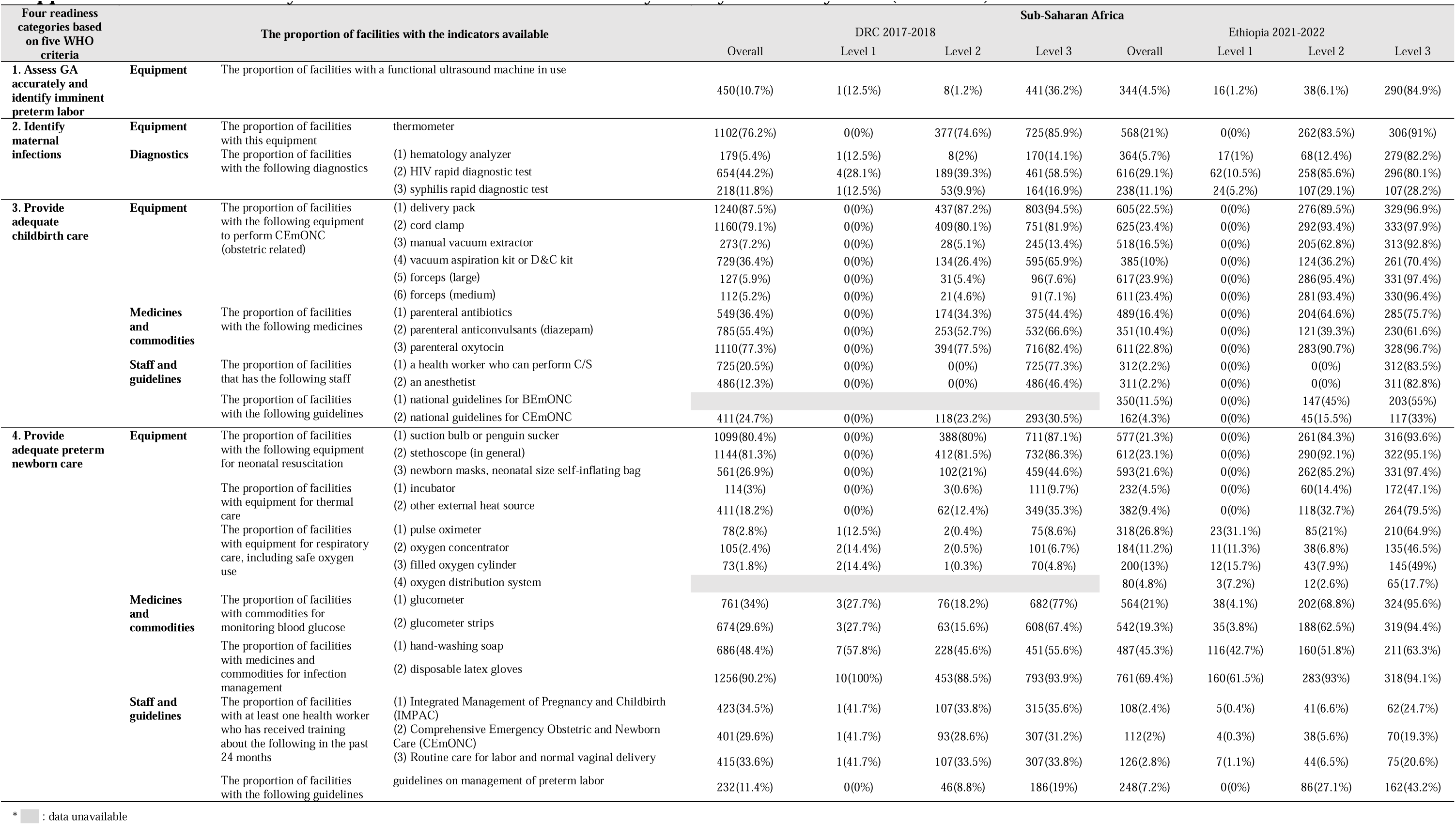
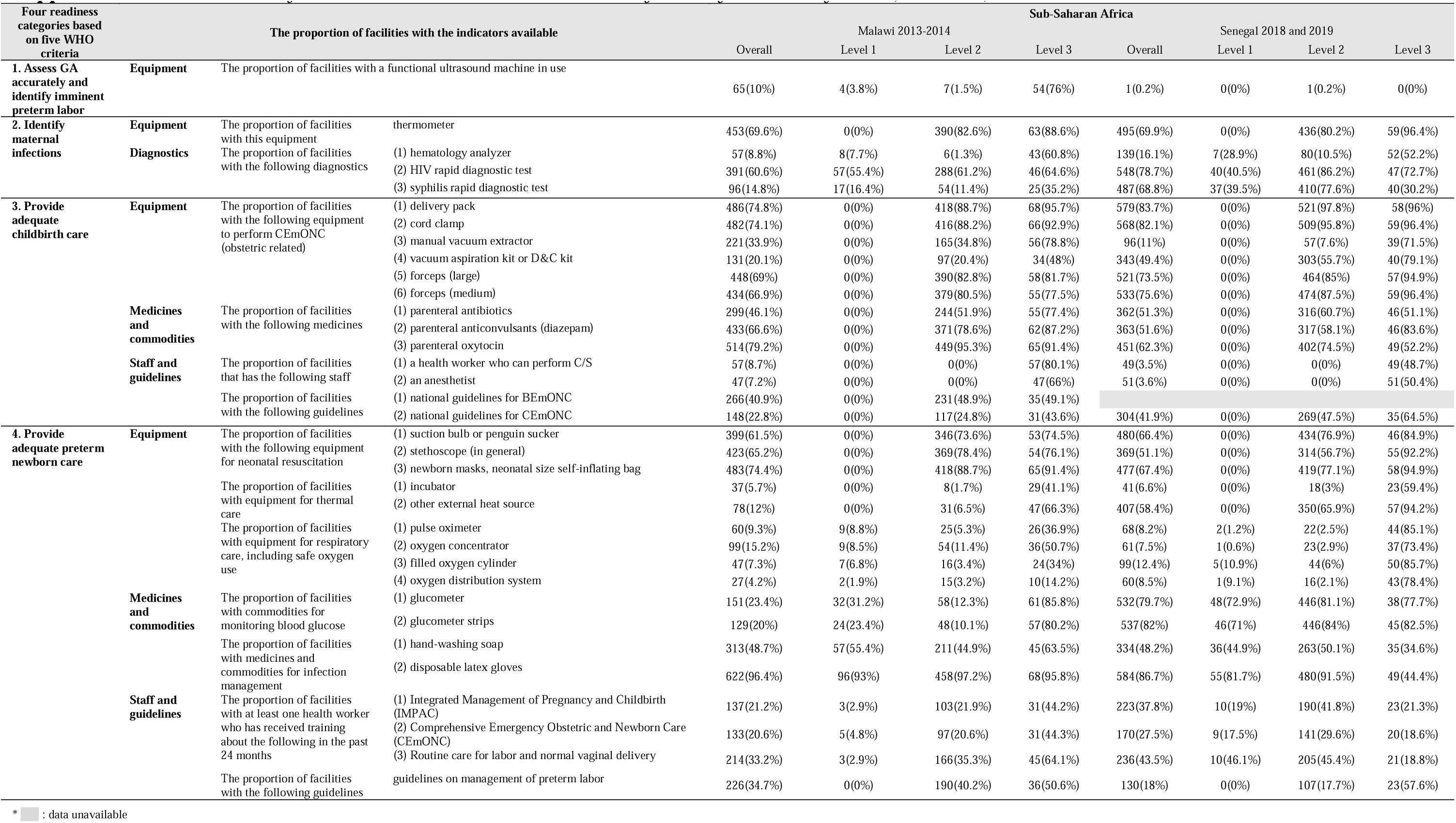
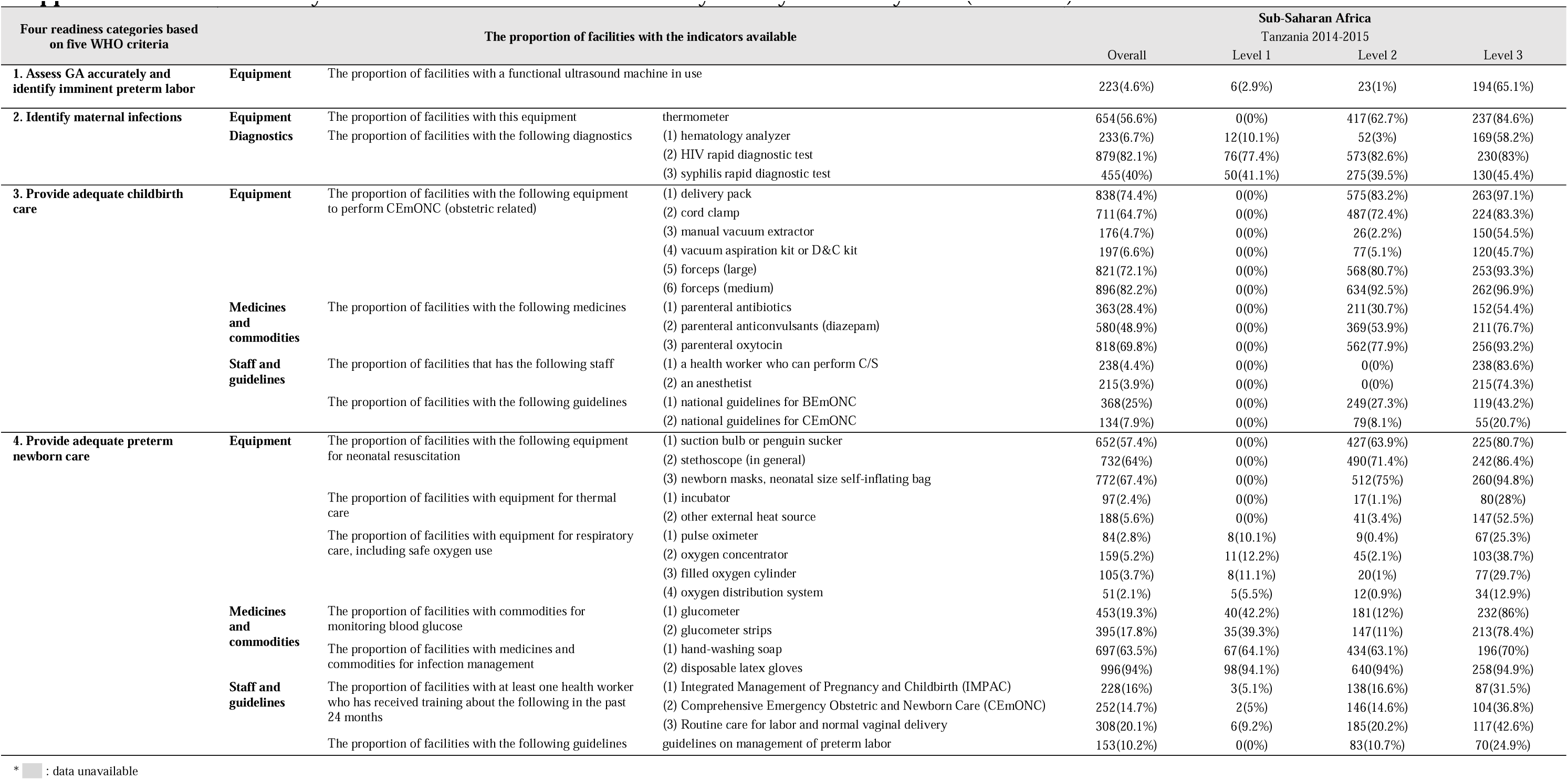
Facility structural readiness for 35 indicators by country and facility level.

**Supplemental Figure 1.**
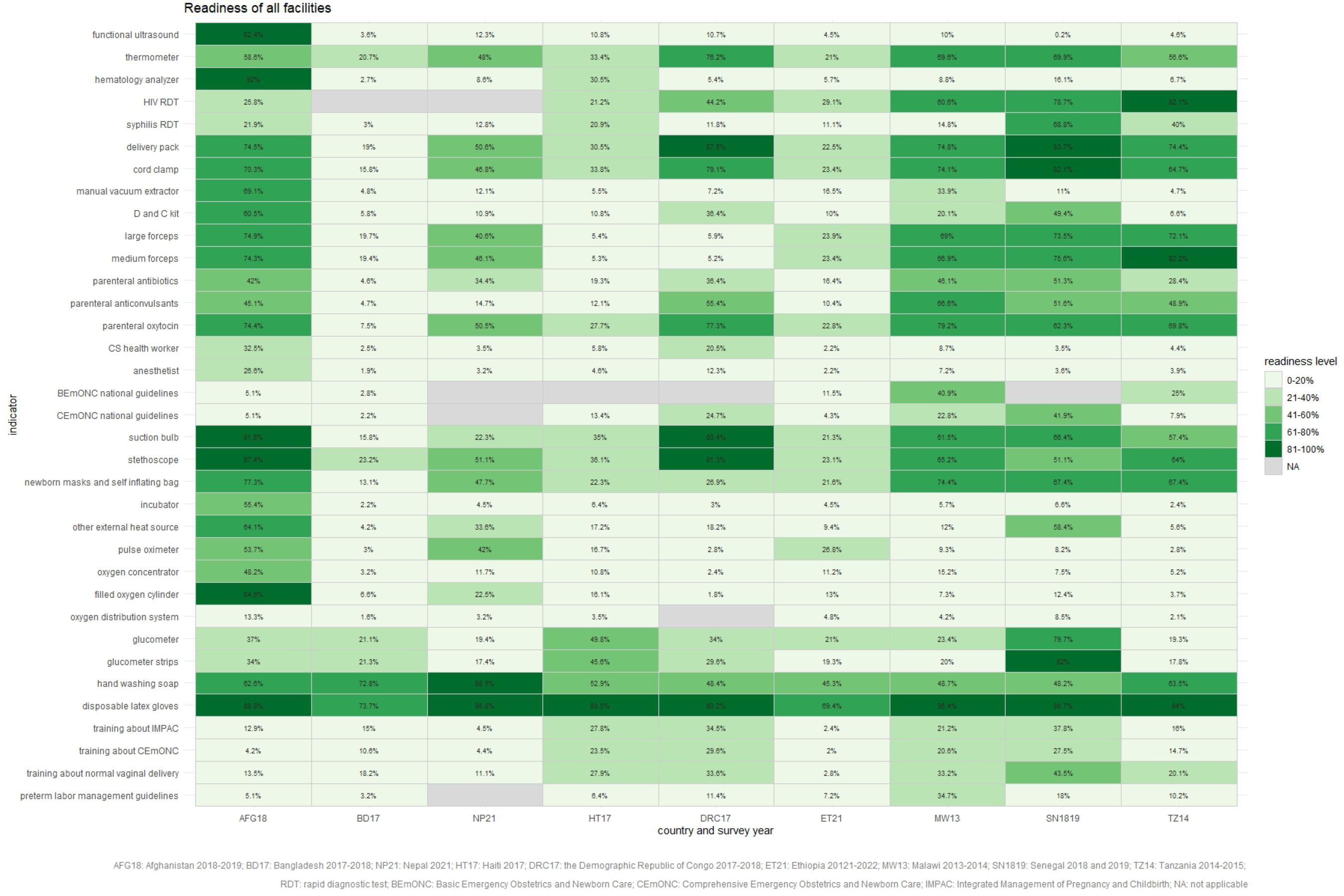
Heatmap of facility readiness for all facilities by country

**Supplemental Figure 2.**
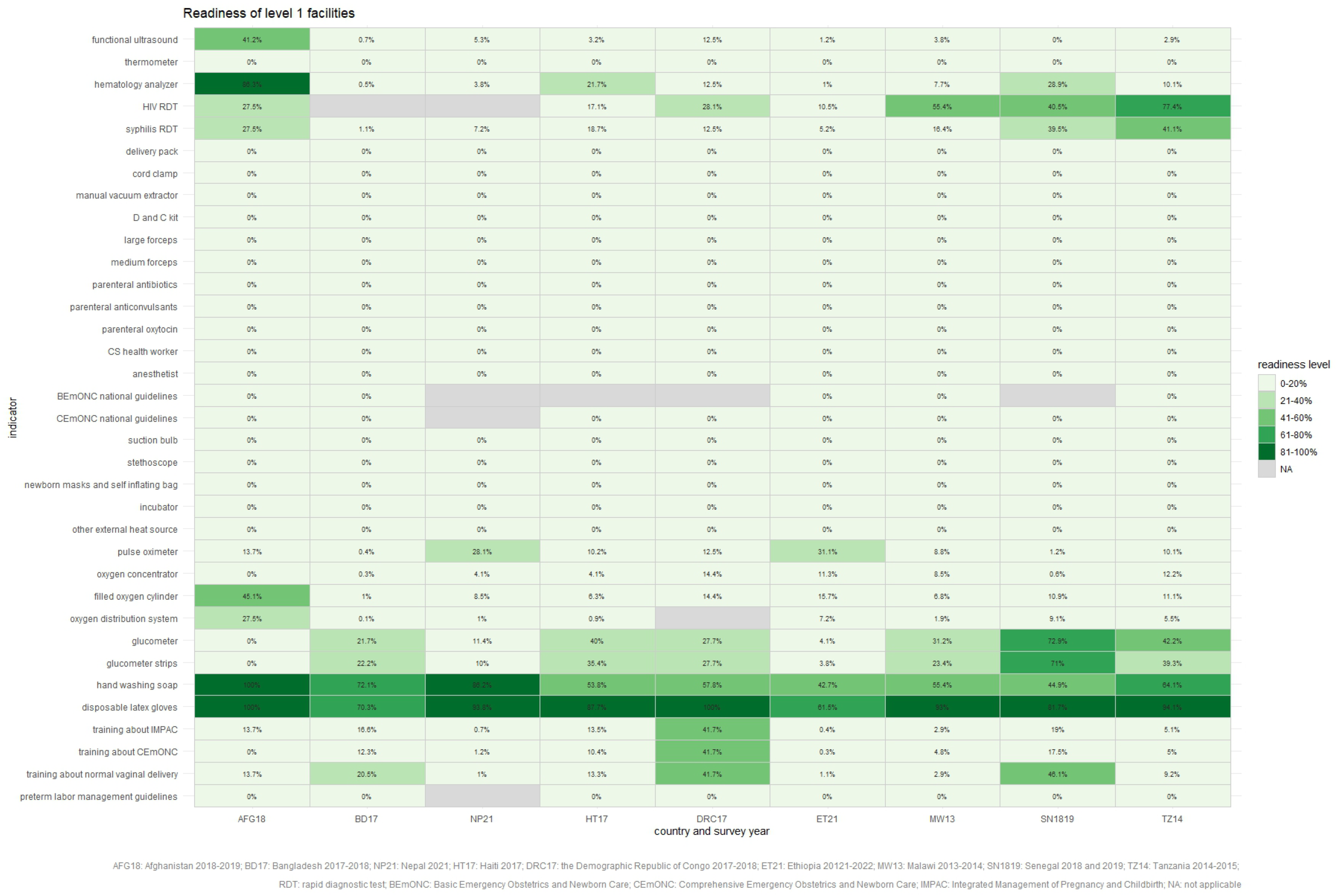
Heatmap of facility readiness for level 1 facilities by country

**Supplemental Figure 3.**
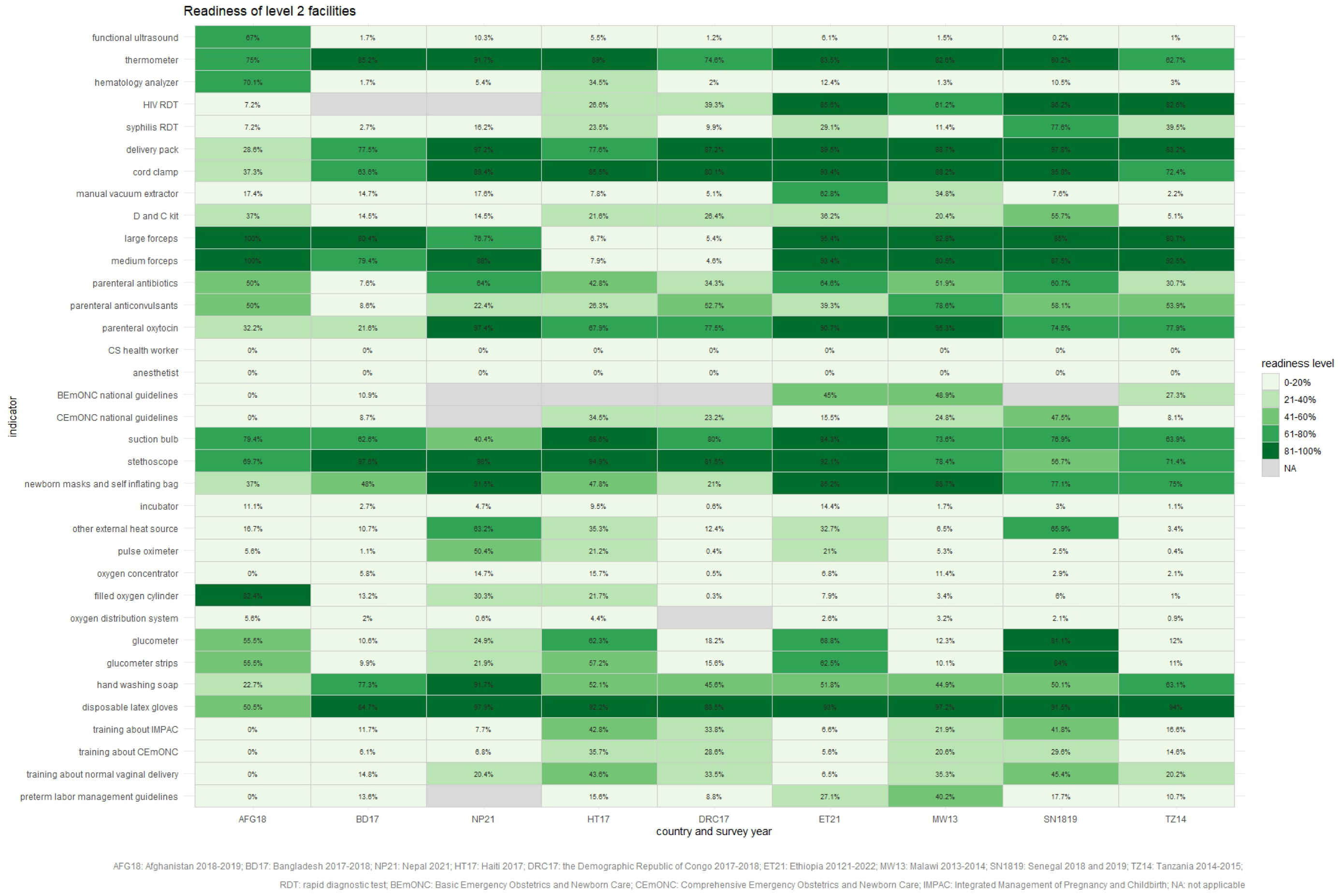
Heatmap of facility readiness for level 2 facilities by country

**Supplemental Figure 4.**
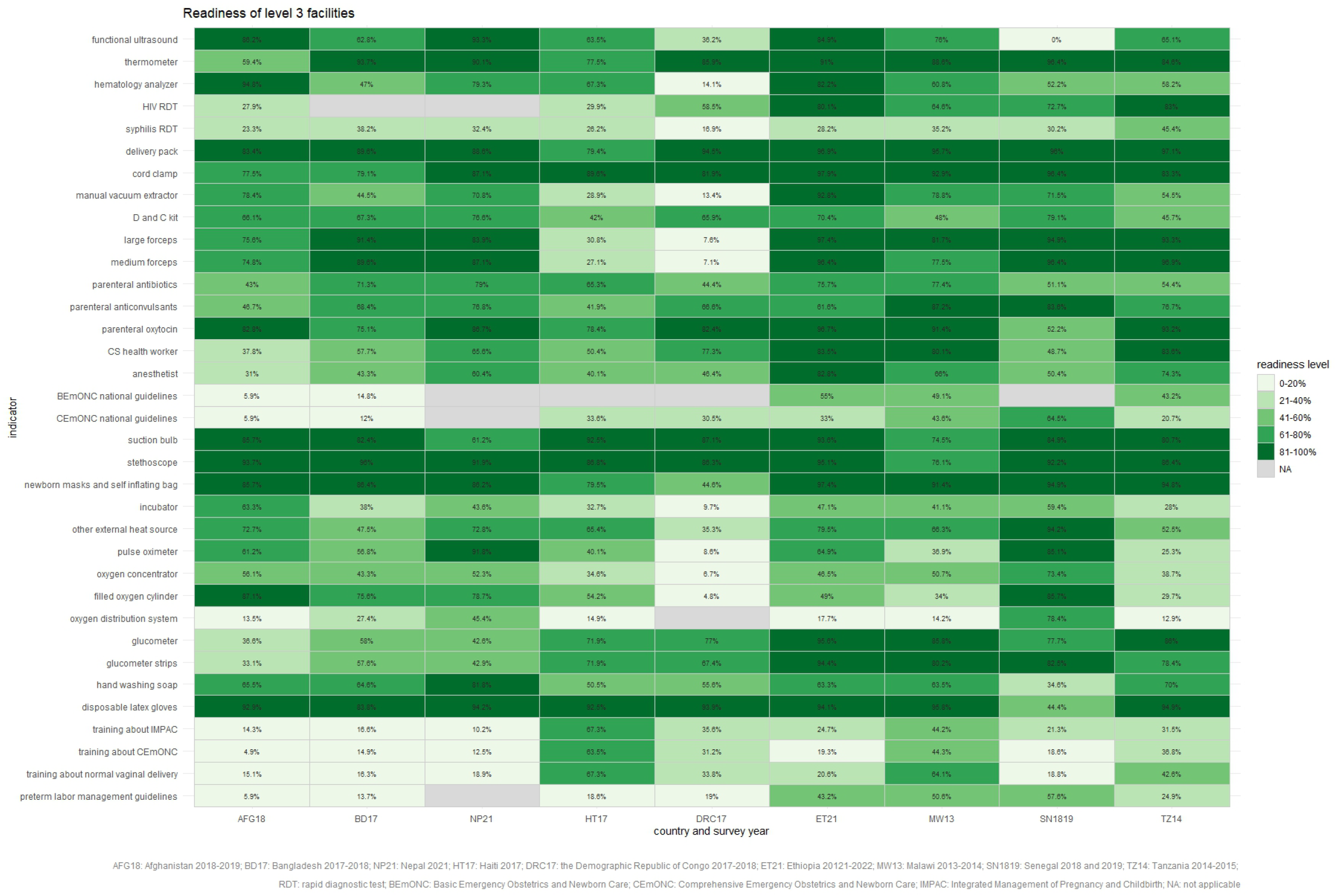
Heatmap of facility readiness for level 3 facilities by country

**Supplemental Figure 5.**
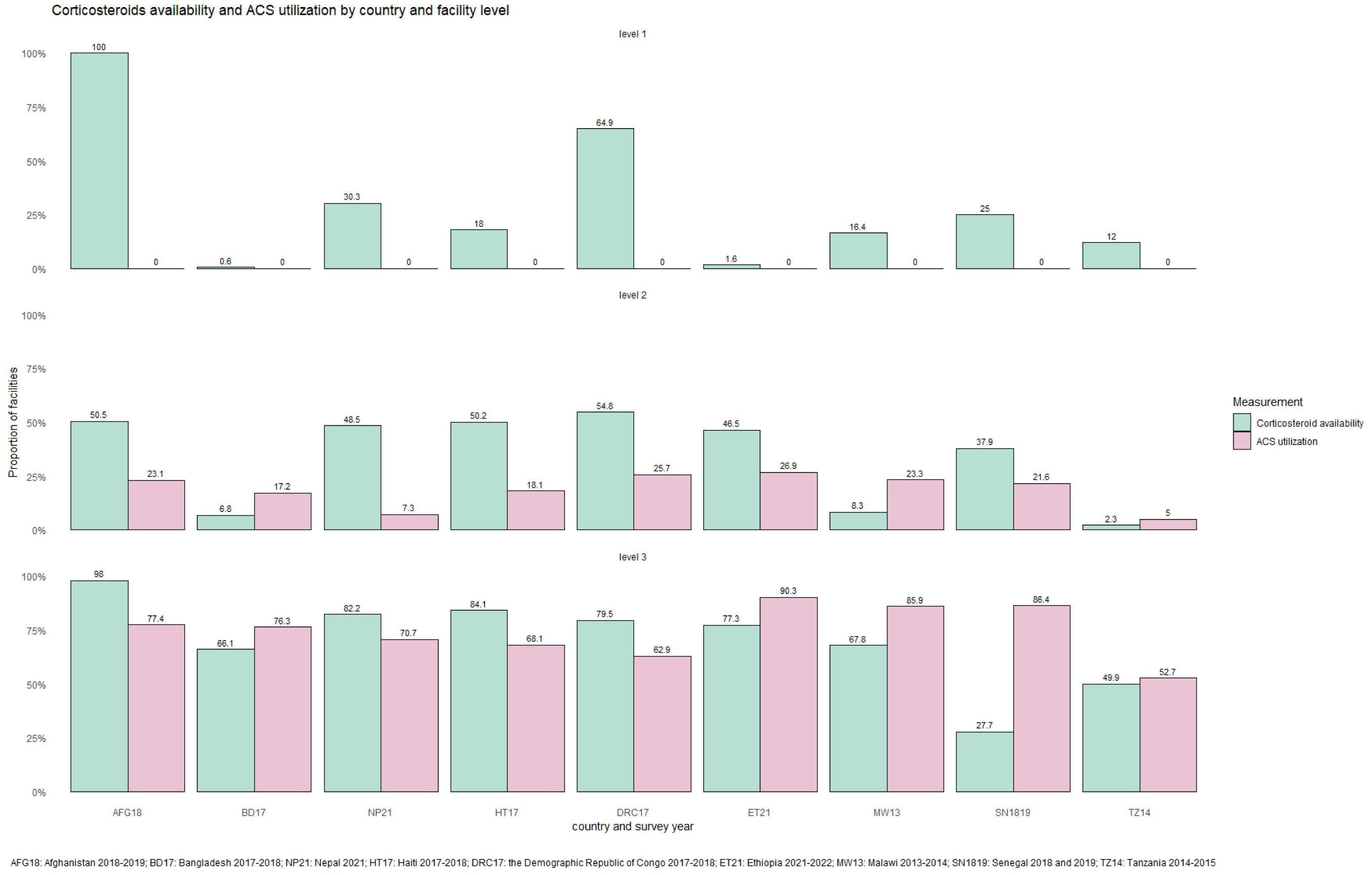
ACS utilization and corticosteroid availability by facility level

**Supplemental Figure 6.**
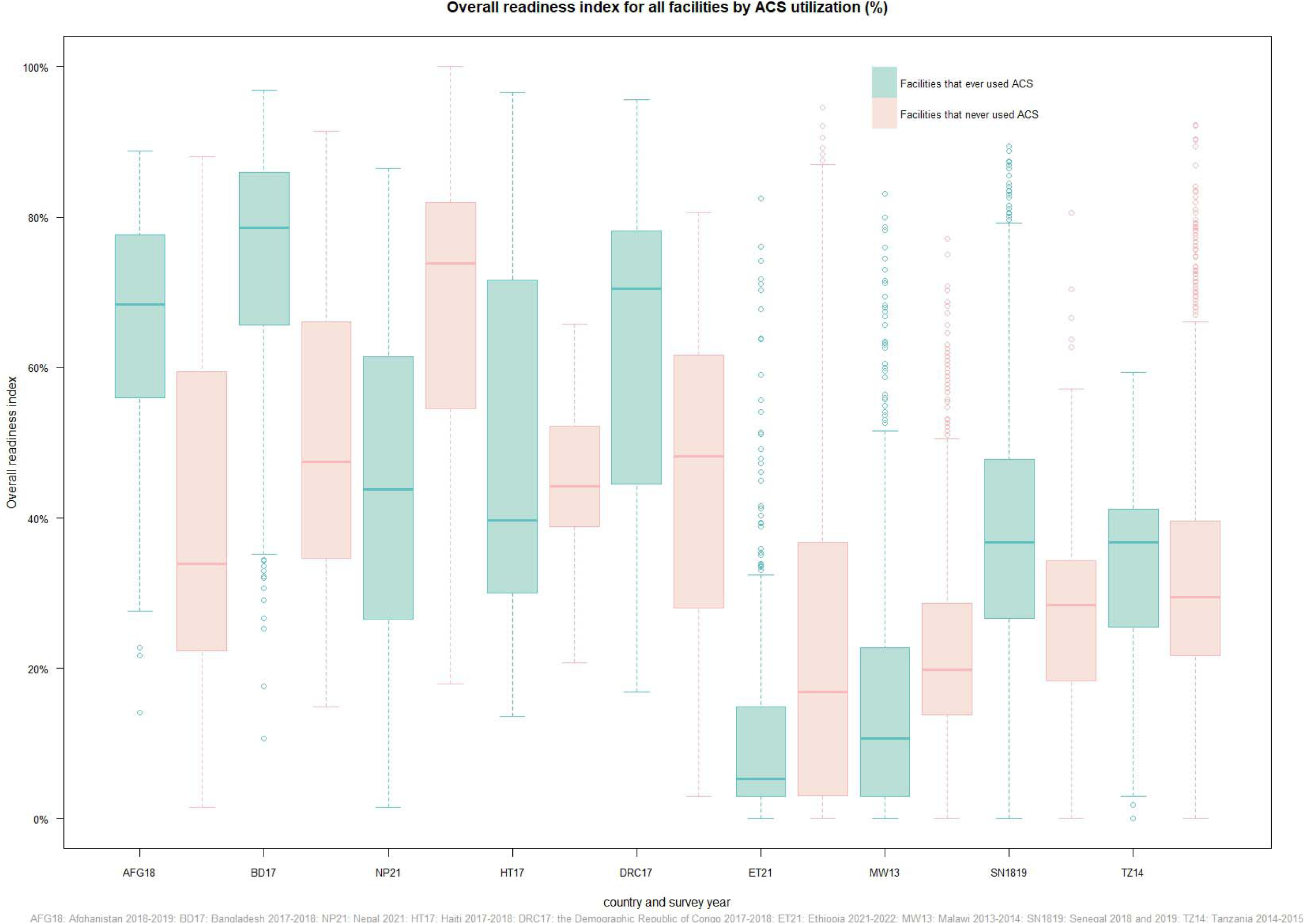
Differences in overall readiness indexes by antenatal corticosteroids utilization for all facilities

**Supplemental Figure 7.**
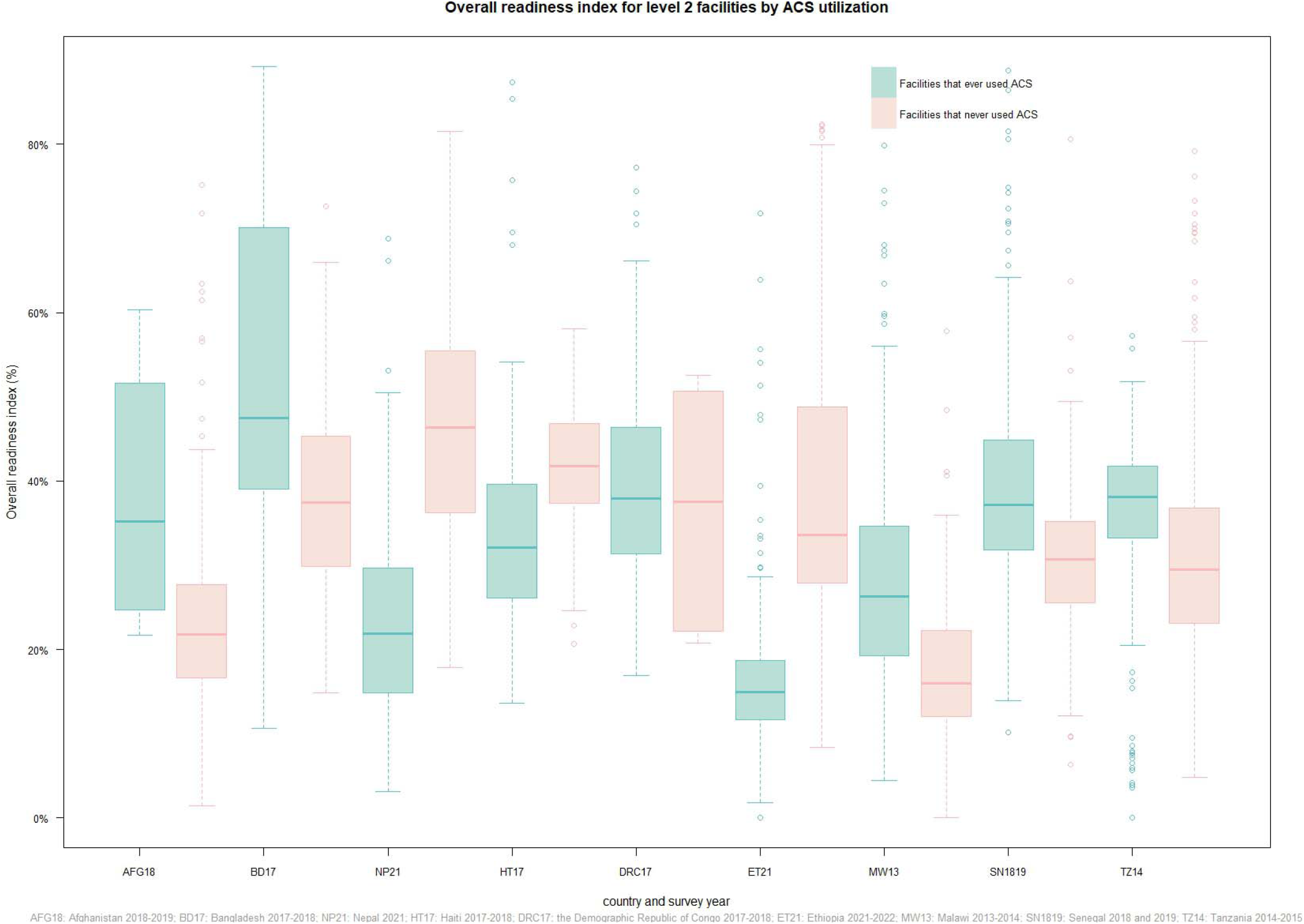
Differences in overall readiness indexes by antenatal corticosteroids utilization for level 2 facilities

**Supplemental Figure 8.**
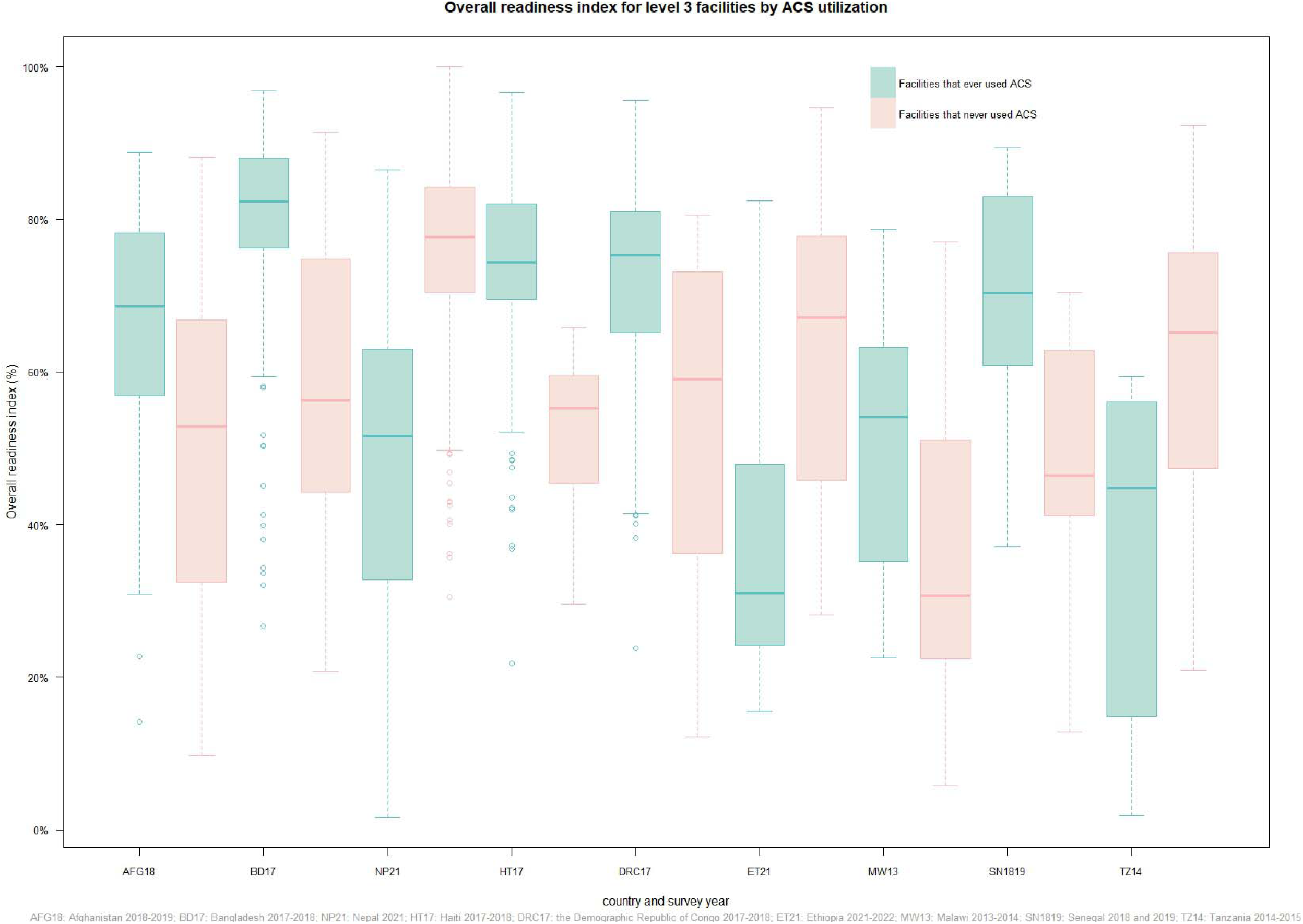
Differences in overall readiness indexes by antenatal corticosteroids utilization for level 3 facilities

